# Bone marrow S-phase is Associated with Risk assessment and Shows Differential Correlation with levels of CD34^+^ Blasts and CD16^−^ Granulocytes in Patients with Myelodysplastic syndromes

**DOI:** 10.1101/2024.06.06.24308422

**Authors:** David Azoulay, Avigail Paz, Galia Stemer

## Abstract

**Introduction:** Including of proliferative parameters in the routine clinical flow-cytometry (FC) workup may be important for better evaluation of prognosis and risk assessment in patients with myelodysplastic syndromes (MDS). However, supportive data are still contradicting.

**Aim:** To assess the levels of FC based proliferative parameters in patients with MDS using propidium Iodide, and to study their correlation with risk assessment and the incidences of CD34^+^ blasts and CD16^-^ and CD16^+^ granulocytes.

**Methods:** Data was retrospectively collected and analysed from 40 specimens of bone marrow (BM) aspiration of MDS patients, 14 specimens of hypocellular BM with no evidence of malignancy, and 16 specimens of reactive BM as determined by pathological examination. IPSS-R scores were calculated for MDS patients then divided to high-risk (IPSS-R score >4.5) and low-risk (IPSS-R score <3). The levels of proliferative parameters and their correlation with incidence of CD34^+^ blasts and CD16^-^ and CD16^+^ granulocytes were compared between high and low risk MDS patients.

**Results:** MDS and reactive BM specimens showed similar whole sample S-phase percentage that was significantly higher than hypocellular BM specimens. As compared to MDS, reactive BM show a greater S-phase percentage in the erythroid fraction. In MDS, the whole sample S-phase percentage was significantly high in a subset of high-risk patients. Increased S-phase percentage in these patients was attributed to high S-phase percentage in granulocytes and monocytes. S-phase percentage correlated with the level of CD34^+^ blasts in the low-risk group but not in the high-risk group. In contrast to CD34^+^ blasts, the S-phase percentage correlated with the level of CD16^-^ granulocytes in the high-risk group but not in the low-risk group.

**Conclusions:** Our observations confirm the association of increased proliferative parameters and adverse prognosis in MDS, and suggest differential proliferation pattern of CD34^+^ and CD16^-^ granulocytes in low and high-risk MDS patients.

## Introduction

Myelodysplastic syndromes (MDS) are a group of clonal myeloid disorders characterized by defective and ineffective haematopoiesis, possibly due to a wide variety of mutations ^1^. The accumulation of mutations contributes to the progression of the disease to Acute Myeloid Leukemia (AML). Consequently, a main measure of the flow-cytometry (FC) in MDS includes monitoring the level of CD34^+^ blasts in the bone marrow (BM) or peripheral blood (PB) to confirm or rule-out transformation to AML ^2,3^. In addition, a detailed characterization of antigenic profile describing the aberrant maturation of the different hematopoietic cell lineages in the BM is another FC based approach for MDS diagnosis and follow-up ^5^. However, new FC based biomarkers are still needed for improving MDS diagnosis and prognosis.

The use of dynamic markers for DNA-cell-cycle analysis in the routine diagnostic FC panel, is a well-established technique which adds an important information on the proliferation and apoptosis of the cells ^8,9^. Our laboratory is experienced in including propidium iodide (PI), a dye that intercalates the DNA and its fluorescence emission is proportional to DNA content in cells, as part of the routine FC diagnostic workup. In this method, FC histograms of DNA in normal cells typically demonstrate a major peak of pre-synthetic G_0_/G_1_ cells, an area corresponding to cells in DNA synthesis (S-phase) and a second peak representing post-synthetic mitotic G_2_M cells. Cell cycle fractions are calculated and the results are given as percentages of cells in G_0_/G_1_, S and G_2_M phases, whereas S and G_2_M could be combined as the proliferative fraction ^10^. Sub G_0_/G_1_ peak represents cells with degraded DNA that corresponds with apoptotic cells ^11^. Extra peaks indicate the presence of tumour cells with abnormal DNA content (DNA aneuploidy). Staining of cells with PI in combination with monoclonal antibody (mAb) against other CDs, such as CD45, enables reliable assessment of cells in various cell cycle phases and evaluation of proliferation and apoptosis in different hematopoietic cell populations ^12^.

Previous studies, demonstrated the usefulness of DNA-cell-cycle analysis by FC in detection of DNA aneuploidy and determination of malignant cell clones ^13^. Another application that is being developed in our laboratory is the measurement of proliferative fraction in lymphocytes as tool that could help in differentiating between aggressive and indolent Non-Hodgkin’s B-cell lymphomas (B-NHLs) ^14^. Descriptive literature regarding the role of proliferative parameters in myeloid neoplasms including MDS are limited showing controversial results and lack of definitive conclusions ^15^. This controversy may be explained by the usage of dyes with dissimilar properties across the studies. For instance, using DRAQ5 in a study by Matarraz et al ^16^, show association between reduced proliferation and shorter survival in 106 MDS patients. In contrast, studies using Ki67 and PCNA in 54 ^17^ and 51 ^18^ MDS patients respectively by Alexandraiks et al, show association between increase proliferation and higher risk of transformation to AML. Of a note, the only study using PI staining of Riccardi et al. ^19^, show association between increase proliferation and shorter survival in 46 AML patients.

Taken together, despite the controversy seen by previous studies, proliferation and apoptotic markers are identified as promising prognostic biomarkers for MDS. Furthermore, the convenience and simplicity of combining dynamic markers for proliferation in the FC diagnostic panel of MDS, identifies it as very attractive tool for investigation. The objective of this study is to assess S-phase percentage in fresh BM aspirations and to define its correlation with the levels of CD34^+^ blasts and CD16^-^ and CD16^+^ granulocytes in patients with high and low-risk MDS.

## Material and Methods

### Case selection

After obtaining an authorization from the Helsinki ethical committee of the Galilee Medical Centre, data on 40 BM specimens with a final diagnosis of MDS that were submitted for evaluation by FC during 2020-2022, were collected in a consecutive order and retrospectively analysed. MDS group included specimens from, 28 males (70%) and 12 females (30%) at an age (mean ± SD) of 71 ± 9.9, median 73 and range 31-93 years. Twenty-nine (72.5%) of the MDS specimens were taken as part of the diagnosis process, the other 11 specimens (27.5%) were on treatment at the time of analysis. The treatments were GCSF in 2 patients, JAK inhibitor in one patient, Venetoclax in one patient, 5-Azacytidine in one patient, Venetoclax + 5-Azacytidine in 6 patients (3 of which with the addition of Dasatinib). Using the IPSS-R scoring system 4, the patients in the MDS group were stratified into low-risk including cases with IPSS-R score less than 3 and high-risk including cases with IPSS-R score greater than 4.5. Cases with intermediate scores were few and did not include in any group. Additional 14 specimens of hypo cellular BM with no evidence of malignancy and 16 specimens of reactive BM were collected as control groups. The hypo cellular BM group included 7 males (50%) and 7 females (50%) at median age of 67 and range 8-83 years. The reactive BM group included 11 males (69%) and 5 females (31%) at median age of 61 and range 15-82 years. Using the IPSS-R scoring system ^4^, the patients in the MDS group were stratified into low-risk including cases with IPSS-R score less than 3 and high-risk including cases with IPSS-R score greater than 4.5. Cases with intermediate scores were not included in any group.

### DNA cell cycle analysis by FC

DNA-cell-cycle analysis by FC was performed on fresh BM aspirates without any preservative using the Coulter DNA prep REAGENT Kit (Ref# 6607055 Beckman Coulter Inc. Brea CA). The estimation of cells in G_0_/G_1_, S and G_2_M compartments in the whole sample or in separated cell subsets according to CD45/SSC gating strategy (co-staining with Pacific-Blue conjugated anti-human CD45 mAb Ref# A74763 Beckman Coulter Inc. Brea CA), were determined by automatic analysis using the ModFit LT software for Windows (version 5.0.9). Additional FC data that we obtained from the specimens included the percentages of CD34^+^ blasts, CD45^+^/ SSC^high^/ CD11b^+^/ CD16^-^ granulocytes and CD45^+^/ SSC^high^/CD11b^+^/CD16^+^ granulocytes.

### Statistical Analysis

Two sided F or t-test were performed to compare the percentage of cells in cell cycle compartments between specimens of MDS, hypo-cellular and reactive BM and between specimens of MDS patients with IPSS-R score >4.5 and IPSS-R score <3. Linear regression test was performed to analyse the correlation between the percentages of cells in S-phase and the percentages of CD34^+^ blasts, CD16^-^ and CD16^+^ granulocytes in low-risk and high-risk MDS. All statistical analyses were performed using JMP (SAS Inc.) statistical software.

## Results

### 1. MDS patients show variable S-phase percentage with mean similar to reactive BM and higher then hypocellular BM

The percentage of cells in subG0/G1 representing the apoptotic cells in the whole sample show no significant differences between MDS, hypo cellular and reactive BM. The S-phase percentage was similar between MDS and reactive BM and significantly higher than hypo cellular BM (*p* = 0.02). The inverse results were seen between MDS, reactive BM and hypo cellular group compering G0/G1 phase (*p* =0.016). (Table 1). The estimations of S-phase percentage in lymphocytes, monocytes, granulocytes and erythroid cells, based on CD45/SSC gating strategy demonstrated significantly higher S-phase percentage in the CD45^**-**^ erythroid cells in the reactive BM compared to MDS (mean S-phase percentage in the CD45^**-**^ erythrocyte fraction ± SD; *21*.*8* ± 12.98 *vs. 48*.*77* ± 12.98 in MDS vs. reactive BM respectively, *p* = 0.02). A trend of higher S-phase percentage in the granulocytes was observed in MDS compared to the reactive and the hypo cellular BM groups (Figure 1).

**Table 1:** Cell cycle fractions in study groups.

| Cell cycle Fraction | MDS<br>% Cells mean<br>± SD<br>median<br>(Range) | Reactive | Hypo cellular | P value |
| --- | --- | --- | --- | --- |
| **sub G0/G1**<br>**(Apoptotic**<br>**cells)** | 10.12 ± 10.22<br>7.04 (0-38.42) | 7.82 ± 7.5<br>5.4 (0-24.4) | 10.12 ± 10.22<br>7.04 (0-38.42) | 0.6 |
| **G0/G1** | 86.16 ± 9.04<br>87.4 (72.8-97.7) | 82.27 ± 5.95<br>88.02 (75.6-98.3) | 93.16 ± 4.1<br>94.4 (86.8-99.23) | 0.016 |
| **S- Phase** | 13.89 ± 9.37<br>12.61 (1.06-38.21) | 12.67 ± 5.9<br>12 (1.56-24.4) | 6.76 ± 4.11<br>5.53 (0.75-13.7) | 0.018 |
| **G2/M** | 0.23 ± 0.84<br>0 (0-5) | 0.05 ± 0.16<br>0 (0-0.63) | 0.12 ± 0.29<br>0 (0-1.06) | 0.34 |

**Figure 1.**
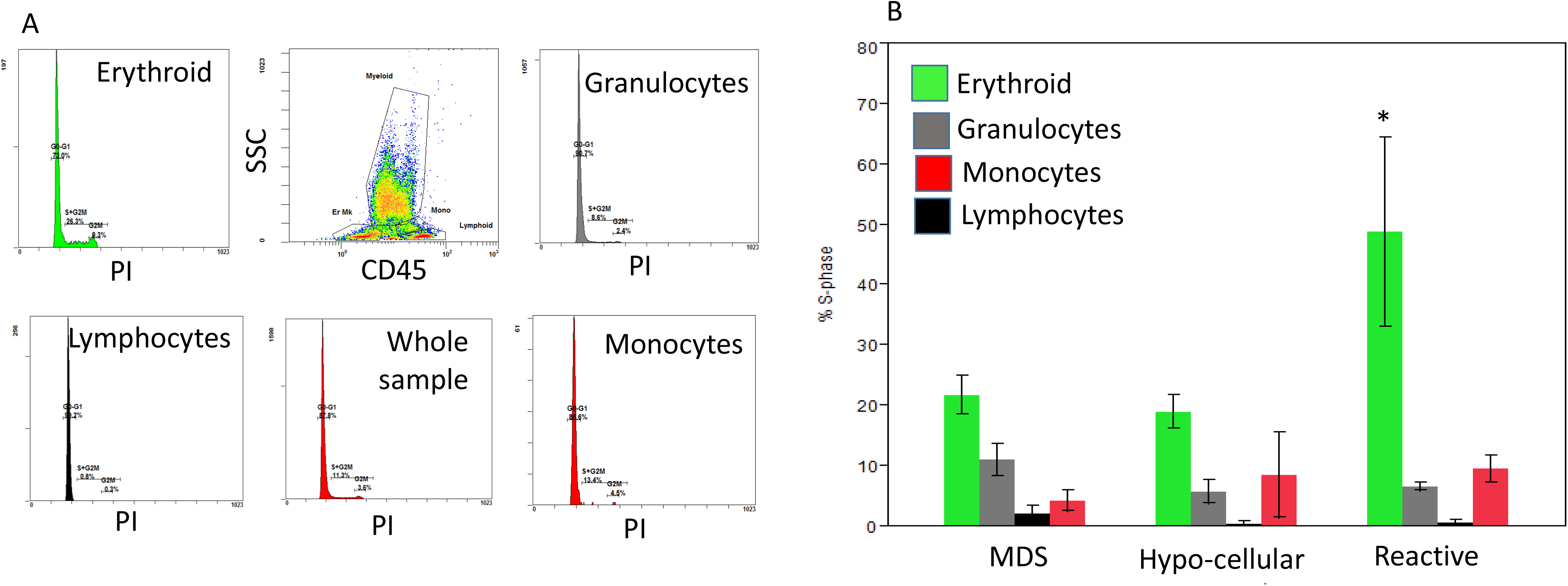
Increased S-phase percentage in the Erythroid cells of Reactive BM specimens. A. plots showing representative histograms of PI-stained BM cells in Erythroid, Myeloid, Monocytes and Lymphocytes based on their CD45/SSC profile. B. Bars showing the mean ± SEM of S-phase percentage in the different subsets between MDS, Hypocellular and Reactive specimens; *\*p* = 0.02.

### 2. S-phase percentage is variable and significantly higher in high-risk MDS patients as compared to low-risk MDS patients

To verify a link between the level of proliferating parameters and the clinical prognosis in MDS, the MDS group was stratified into high and low risk groups according to the IPSS-R score. IPSS-R was completed in 25 patients. High-risk group included all patients with IPSS-R score 4.5 or greater and low-risk included all patients with IPSS-R 3 or lower. The high-risk group showed high variability of S-phase percentage and overall significantly higher percentage of cells in S-phase percentage compared to low-risk group (18.45 ± 11.43 vs. 8.39 ± 4.69 in high vs. low-risk MDS patients group respectively, p = 0.02) (Figure 2A). One patient with intermediate score, showed S-phase percentage similar to the high-risk group (not shown). A search for a sub-populations of cells contributing to the increase of whole sample S-phase percentage in the high-risk patients, revealed significantly higher S-phase percentage in granulocytes (mean S-phase percentage ± SD; *29*.*9* ± 8.85 *vs. 3*.*73* ± 3.07 in high-risk patients with a whole sample S-phase percentage > 20% vs. < 20% respectively, p<0.001) and in monocytes (mean S-phase percentage ± SD; *17*.*14* ± 10.22 *vs. 1*.*31* ± 2.62 in high-risk patients with whole sample S-phase percentage ≥ 20% vs. < 20% respectively, p<0.03). (Figure 2B). Notably, no other significant differences in the proportion of patients under treatment and nor type of cytogenetic aberrations or mutations were found between high-risk patients with whole sample S-phase percentage ≥ 20% vs. < 20%.

**Figure 2.**
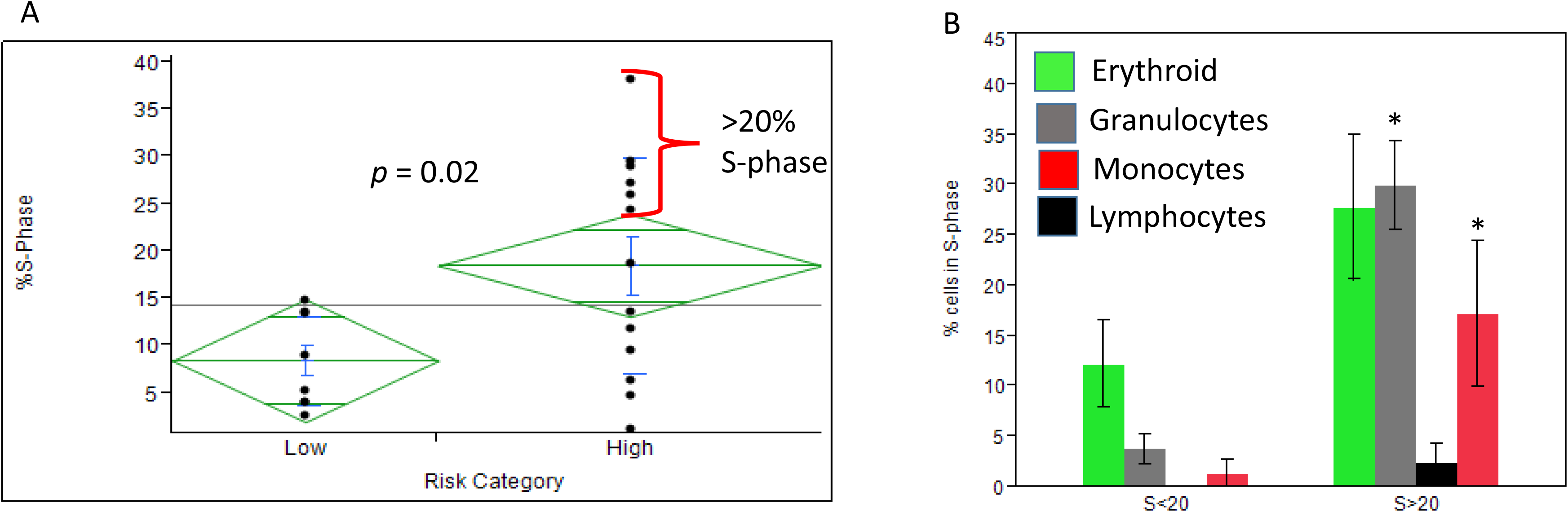
Increased S-phase percentage in high-risk MDS patients. A. plot showing the variability and the significantly increased levels of S-phase percentage in High risk MDS patients as compared to low risk MDS patients; *\*p* = 0.02. The subset of high risk MDS patients with >20% S-phase are marked with the red marker. B. Bars showing the mean ± SEM of S-phase percentages in Erythroid, Myeloid, Monocytes and Lymphocytes between high-risk MDS with >20% S-phase and <20% S-phase; *\*p* < 0.05.

### 3. The level of cells in S-phase percentage correlated with the level of CD34^+^ cells in low-risk MDS and with CD16^-^ and CD16^+^ granulocytes in high-risk MDS

We checked differences in the correlation between S-phase percentage and the levels of CD34^+^ cells and the CD16^-^ and CD16^+^ granulocytes as determined by the FC in the specimens between low and high-risk MDS patients. A significant positive correlation was found between the whole sample S-phase percentage and CD34^+^ cells in the low-risk group (*r*=0.83, p<0.0001) but not in the high-risk group (*r*=0.26, p=0.37). In contrast, the whole sample S-phase percentage shows significant positive correlation with the level of CD16^-^ granulocytes in the high-risk group (*r = 0*.*85, p<0*.*0001)* and not in the low-risk group (*r = 0*.*65, p=0*.*108)*. Both low and high-risk MDS patients showed negative correlation between S-phase and CD16^+^ granulocytes (Table 2).

**Table 2:** Correlations between S-phase percentage, CD34^+^ blasts, CD16^-^ and CD16^+^ granulocytes between low and high risk MDS.

| Correlation | MDS Low-risk | MDS High-risk |
| --- | --- | --- |
| S- Phase vs.<br>CD34<sup>+</sup> Blasts | 0.83, <0.0001 | 0.26, 0.37 |
| S- Phase vs.<br>CD16<sup>-</sup><br>granulocytes | 0.65, 0.108 | 0.87, <0.0001 |
| S- Phase vs.<br>CD16<sup>+</sup><br>granulocytes | -0.77, 0.04 | -0.8, 0.0095 |

## Discussion

In this study we aimed to assess proliferation parameters in BM specimens of MDS patients using FC based DNA-cell-cycle-analysis with PI staining. The differences in whole sample S-phase percentage between hypo-cellular BM and MDS /reactive BM are coherent and confirm the reproducibility of our assay to determine the proliferative fractions in the specimens. The significantly higher S-phase percentage in the erythroid lineage in reactive BM is in line with previous study (Mestrum *et al*, 2021b), suggesting a more common physiologic reaction in reactive specimens as opposed to a more variable and complexed proliferative changes in MDS patients.

Our observations show high variability of S-phase percentage in MDS patients with high-risk and a subset of high-risk patient’s that present with extremely high S-phase percentage compared to low-risk MDS patients. These findings support previous studies concluding the association between increased proliferation parameters and adverse prognosis in MDS ^17, 18^. However, as indicated by our data, MDS patients with high-risk could also present with low S-phase percentage. Furthermore, as was shown in other studies ^16, 21^, we could not rule out the possibility that using a different DNA dye in our study would have resulted with different trend.

Our attempts to characterize unique pattern in the cytogenetic and the mutational status of high-risk MDS patients with the extremely high S-phase percentage revealed no significant trend. However, our findings suggest that in high-risk MDS patients the pattern of increased proliferative parameters tend to involve a multilineage proliferation rather than a unilineage myeloid proliferation. Whether and how this proliferative feature is linked and promote multilineage dysplastic leukogenesis in the disease ^16,22^ should be further studied.

Finally, analysis of the correlations between S-phase percentage, CD34^+^ blasts and CD16^-^ and CD16^+^ granulocytes between MDS low and high risk, revealed intriguing observations in our study. When the analysis between S-phase percentage and CD34^+^ blasts was performed in all the MDS patients no significant correlation was found. However, when the analysis between S-phase percentage and CD34^+^ blasts was performed separately in low and high-risk MDS patients, a strong positive correlation was observed only in the low-risk MDS patients. This observation suggests that CD34^+^ blasts may proliferate differently or contribute differently to the proliferating cell pool in low-risk MDS compared to high-risk MDS patients. This distinction gained further support from the analysis of the correlations between S-phase percentage and CD16^-^ granulocytes which showed an opposite result. As CD16^-^ granulocytes correspond with immature or dysplastic granulocytes in MDS, these observations are reasonable as myeloid cell proliferation is usually involved the granulocytic lineage in high risk patients but not in low risk patients. Furthermore, these observations suggest that as part of the advanced pathophysiology of the disease, cell proliferation in high risk patients may occurs also in cells with more mature state then the CD34^+^ blasts or in blasts that are lacking CD34. While this possible interpretation is supported by other studies ^16, 20^, a verification to this distinction as an objective pathophysiological future in the advanced disease stages requires more investigation.

This study has several limitations. First, the overall sample size is relatively small and lack of specimens of patients with myeloproliferative disease as controls which could add additional insight to the study. In addition, although most of the MDS patients in the study were at diagnosis some patients were under treatment which could have an effect on the results. Another point is that in our study, FC defined the percentage of blasts by gating CD34^+^ cells. The percentage of blasts in fact, could be underestimated by FC because of CD34^-^ blasts and peripheral blood contamination. Hence, it would be better to combine together FC and morphologic analysis for the percentage of blasts as well as for the CD16^-^ and CD16^+^ granulocytes.

In summary, our study shows differences in the levels of proliferating cells between BM specimens of hypocellular, reactive and MDS patients. The increased levels of S-phase percentage in a subset of patients with high-risk MDS, suggest the association of increased proliferation and adverse prognosis in MDS. The differential correlation patterns of S-phase percentage with CD34^+^ blasts and CD16^-^ granulocytes between low and high-risk MDS may be linked with the disease progression. However, further research is needed to confirm these observations.

## Data Availability

All data produced in the present study are available upon reasonable request to the authors

